# Combination of Antibody based rapid diagnostic tests used in an algorithm may improve their performance in SARS CoV-2 diagnosis

**DOI:** 10.1101/2020.06.26.20140806

**Authors:** Grace Esther Kushemererwa, Ismail Kayongo, Patrick Semanda, Hellen Nansumba, Iga Tadeo, Christine Namulindwa, Wilson Nyegenye, Charles Kiyaga, Patrick Ogwok, Susan Ndidde, Isaac Ssewanyana

## Abstract

**Background:** Globally response to the SARS-CoV-2 pandemic is highly limited by diagnostic methods. Currently, World Health Organization (WHO) recommends the use of molecular assays for confirmation of SARS-CoV-2 infection which are highly expensive and require specialized laboratory equipment. This is a limitation in mass testing and in low resource settings. SARS CoV-2 IgG/IgM antibody tests have had poor diagnostic performance that do not guarantee their use in diagnostics. In this study we demonstrate a concept of using a combination of RDTs in an algorithm to improve their performance for diagnostics.

**Method:** Eighty six (86) EDTA whole blood samples were collected from SARS-CoV-2 positive cases admitted at Masaka and Mbarara Regional Referral Hospitals in Uganda. These were categorized from day when confirmed positive as follows; category A (0-3 days, 10 samples), category B (4-7 days, 20 samples), Category C (8-17 days, 11 samples) and Category D (18-28 days, 20 samples). Plasma was prepared, transported to the testing laboratory and stored at −20^0^C prior to testing. A total of 13 RDTS were tested following manufacturer’s instructions. Data was entered in Microsoft Excel exported to STATA for computation of sensitivity and specificity. We computed for all possible combinations of 2 of the 13 RDTS (13C_2)_ that were evaluated in parallel algorithm.

**Results:** The individual sensitives of the RDTs ranged between 74% and 18% and there was a general increasing trend across the categories with days since PCR confirmation. A total of 78 possible combinations of the RDTs to be used in parallel was computated. The combinations of the 2 RDTS improved the sensitivities to 90%.

**Discussion:** We demonstrate that use of RDTs in combinations can improve their overall sensitivity. This approach when used on a wider range of combination of RDTs may yield combinations that can give sensitivities that are of diagnostics relevance in mass testing and low resource setting.

## Background

The Severe acute respiratory syndrome coronavirus 2 (SARS-CoV-2) is a novel coronavirus that causes coronavirus disease 2019 (COVID-19), the respiratory illness responsible for the COVID-19 pandemic(1). SARS-CoV-2 specific antibodies are produced over days to weeks after infection with the virus. The strength of antibody response depends on several factors, including age, nutritional status, severity of disease, and certain medications or infections like HIV that suppress the immune system(2). Antibody responses have been demonstrated in SARS-CoV-2 with a prevalence of 40% of patients in the first 7 days of illness, and then rapidly increased to 100.0%, 94.3% and 79.8% for Total Ab, IgM and IgG respectively since day 15 after onset(3). Evidence of early acquisition of SARS CoV-2 antibodies strengthen the idea of use of RDTs for mid, late, convalescent, and post-recovery phases of disease.

Rapid diagnostic tests do not require any testing equipment, may be stored at room temperature are easy to operate and are cheap. They also have rapid screening time of approximately 10-15 minutes. These attributes make RDTS a very attractive for mass testing.

Because of the poor performance of the currently available SARS COV-2 antibody RDTs, World Health Organization (WHO) recommends the use of RDT tests only in research and surveillance settings. In our study, we aimed at using 2 RDTS in a test algorithm to determine if it can improve their performance in terms of sensitivity and specificity.

## Methodology

### Sample size and characterization

In a prospective study, a total of 86 samples were tested on each COVID-19 rapid diagnostic kit. Negative samples were retrieved from sample archives in the January 2018 inventory at the National Biorepository. Positive samples were obtained from confirmed positive patients by GeneXpert admitted at Masaka and Mbarara Regional Referral Hospitals and these were categorized from day when confirmed positive as follows; category A (0-3 days) 10 samples, category B (4-7 days) 20 samples, Category C (8-17 days) 11 samples and Category D (18-28 days) 20 samples.

### Specimen collection, transport and storage

Whole blood was collected in commercially available anti-coagulant EDTA tubes. The samples were transported to CPHL for testing as per CPHL standard operating procedures. Plasma was separated, aliquoted into 4 vials and stored according to CPHL standard operating procedures

### Testing

In this study we included 13 RDTs that were provided by different Laboratory suppliers who wanted to have their product evaluated. Testing of the Rapid Diagnostic test kits was carried out at the Central Public health laboratories (CPHL). This is a lateral flow immune chromatographic assay. All the 13 COVID-19 kits use anti human IgM antibody (test line IgM), anti-human IgG (test Line IgG) and IgG (control line) immobilized on a nitrocellulose strip. The burgundy colored conjugate pad contains colloidal gold conjugated to recombinant COVID-19 antigen conjugated with colloid gold (COVID-19 conjugate). When a specimen followed by a buffer is added, IgM and or IgG if present will bind to COVID-19 conjugates making antigen antibody complex. This complex migrates through nitrocellulose membrane by capillary action.

A total of 13 rapid diagnostic kits were verified. 10 μl aliquot of plasma was used on each of the rapid diagnostic test kits and results read between 10-20 minutes following each manufacturer’s instructions on the Kit inserts. Results were recorded on a paper-based study record form (SRF), dated and initiated by a laboratory technologist and reviewed by another laboratory technologist

### Data Management and Analysis

All data was entered into Microsoft excel from the paper-based study record form (SRF). A 2 by 2 table was used to compute for sensitivity, specificity, negative and positive predictive values. 13C_2_ was used to assemble the 78 possible combinations of the Rapid Diagnostic kits to be used in parallel with improved sensitivity.

## Results

All the RDTs have a sensitivity less than 85%. All the RDTs showed increased sensitivity over time, from the day of confirmed SARS CoV-19 infection. All the RDTs had a sensitivity less than 85% by Day (0-3), Day (4-7), and Day (8-14). Only 3 out of 13 RDTs had a sensitivity above 85% by Day (15-18). 12 out of 13 RDTs have a specificity greater than 85%.13 RDTs that were included into this evaluation. A simulated combination of at least 2 RDTs tested in parallel, resulted in 78 combinations. Each of the RDT has an equal chance of combination of the other. A Positive result was scored if either of the RDT within the combination has Positive IgG or IgM test result. And a Negative test result is scored if all of the RDT within the combination had Negative IgG or IgG. Combined Sensitivity was computed as shown in Table 3 below Two (2) Combinations of RDTs with sensitivities above 85% and 100% specificity tested in parallel were selected.

**Table 1:** A Table showing the sensitivity, specificity and 95% CI of each of the 13 IgG / IgM antibody tests.

| Assay ID | Sensitivity | [95% CI ] | Specificity | [95% CI] |
| --- | --- | --- | --- | --- |
| A | 74% | [61%- 84%] | 92% | [74%-99%] |
| B | 74% | [53%-78%] | 72% | [55%-91%] |
| C | 66% | [52%-77%] | 91% | [78%-98%] |
| D | 64% | [51%-76%] | 92% | [74%-99%] |
| E | 59% | [46%-71%] | 97% | [90%-99%] |
| F | 59% | [46%-71%] | 80% | [59%-93%] |
| G | 56% | [39%-65%] | 92% | [74%-99%] |
| H | 49% | [36%-62%] | 92% | [74%-99%] |
| I | 43% | [31%-57%] | 100% | [87%-98%] |
| J | 39% | [27%-53%] | 96% | [80%-100%] |
| K | 41% | [29%-54%] | 80% | [59%-93%] |
| L | 37% | [25%-50%] | 96% | [80%-100%] |
| M | 18% | [8%-27%] | 100% | [86%-100%] |

**Table 2:** A table showing the sensitivity of each the 13 IgG / IgM antibody tests at Day (0- 3), Day (4-7), Day (8-14) and Day (15-28)

| Assay ID | Sensitivity |  |  |  |
| --- | --- | --- | --- | --- |
|  | Day 0-3 | Day 4-7 | Day 8-14 | Day 15-28 |
| A | 70% | 70% | 73% | 80% |
| B | 80% | 65% | 64% | 90% |
| C | 50% | 65% | 45% | 85% |
| D | 60% | 55% | 55% | 80% |
| E | 44% | 55% | 36% | 90% |
| F | 40% | 50% | 55% | 80% |
| G | 20% | 55% | 45% | 70% |
| H | 30% | 45% | 36% | 70% |
| I | 40% | 55% | 55% | 63% |
| J | 10% | 35% | 36% | 65% |
| K | 30% | 40% | 27% | 55% |
| L | 10% | 25% | 40% | 60% |
| M | 0% | 20% | 0% | 35% |

**Table 2:** A table showing proposed algorithm to simulate combination of at least 2 RDTS tested in parallel.

| No. | Common name |  |  |  |  |  |  |  |  |  |  |  |  |  | Total |
| --- | --- | --- | --- | --- | --- | --- | --- | --- | --- | --- | --- | --- | --- | --- | --- |
| 1 | A | Combinations of 1 | 1, 2 | 1, 3 | 1, 4 | 1, 5 | 1, 6 | 1, 7 | 1, 8 | 1, 9 | 1, 10 | 1, 11 | 1, 12 | 1, 13 | 12 |
| 2 | B | Combinations of 2 |  | 2, 3 | 2, 4 | 2, 5 | 2, 6 | 2, 7 | 2, 8 | 2, 9 | 2, 10 | 2, 11 | 2, 12 | 2, 13 | 11 |
| 3 | C | Combinations of 3 |  |  | 3, 4 | 3, 5 | 3, 6 | 3, 7 | 3, 8 | 3, 9 | 3, 10 | 3, 11 | 3, 12 | 3, 13 | 10 |
| 4 | D | Combinations of 4 |  |  |  | 4, 5 | 4, 6 | 4, 7 | 4, 8 | 4, 9 | 4, 10 | 4, 11 | 4, 12 | 4, 13 | 9 |
| 5 | E | Combinations of 5 |  |  |  |  | 5, 6 | 5, 7 | 5, 8 | 5, 9 | 5, 10 | 5, 11 | 5, 12 | 5, 13 | 8 |
| 6 | F | Combinations of 6 |  |  |  |  |  | 6, 7 | 6, 8 | 6, 9 | 6, 10 | 6, 11 | 6, 12 | 6, 13 | 7 |
| 7 | G | Combinations of 7 |  |  |  |  |  |  | 7, 8 | 7, 9 | 7, 10 | 7, 11 | 7, 12 | 7, 13 | 6 |
| 8 | H | Combinations of 8 |  |  |  |  |  |  |  | 8, 9 | 8, 10 | 8, 11 | 8, 12 | 8, 13 | 5 |
| 9 | I | Combinations of 9 |  |  |  |  |  |  |  |  | 9, 10 | 9, 11 | 9, 12 | 9, 13 | 4 |
| 10 | J | Combinations of 10 |  |  |  |  |  |  |  |  |  | 10, 11 | 10, 12 | 10, 13 | 3 |
| 11 | K | Combinations of 11 |  |  |  |  |  |  |  |  |  |  | 11, 12 | 11, 13 | 2 |
| 12 | L | Combinations of 12 |  |  |  |  |  |  |  |  |  |  |  | 12, 13 | 1 |
| 13 | M | Combinations of 13 |  |  |  |  |  |  |  |  |  |  |  |  | 0 |
|  |  |  |  |  |  |  |  |  |  |  |  |  |  |  | 78 |

**Table 3:**
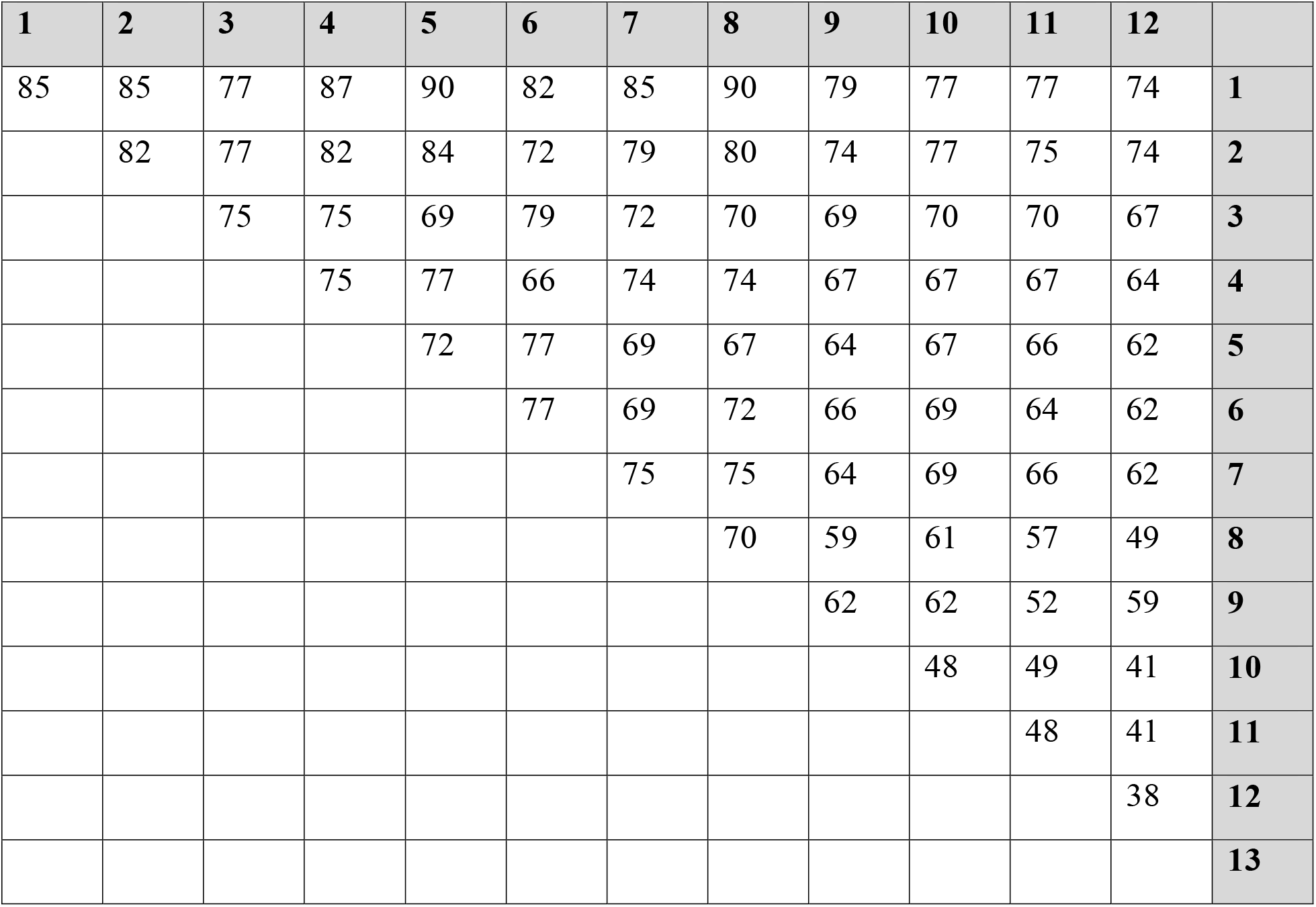
Table showing computed sensitives from the 78 combinations.

| **1** | **2** | **3** | **4** | **5** | **6** | **7** | **8** | **9** | **10** | **11** | **12** |  |
| --- | --- | --- | --- | --- | --- | --- | --- | --- | --- | --- | --- | --- |
| 85 | 85 | 77 | 87 | 90 | 82 | 85 | 90 | 79 | 77 | 77 | 74 | **1** |
|  | 82 | 77 | 82 | 84 | 72 | 79 | 80 | 74 | 77 | 75 | 74 | **2** |
|  |  | 75 | 75 | 69 | 79 | 72 | 70 | 69 | 70 | 70 | 67 | **3** |
|  |  |  | 75 | 77 | 66 | 74 | 74 | 67 | 67 | 67 | 64 | **4** |
|  |  |  |  | 72 | 77 | 69 | 67 | 64 | 67 | 66 | 62 | **5** |
|  |  |  |  |  | 77 | 69 | 72 | 66 | 69 | 64 | 62 | **6** |
|  |  |  |  |  |  | 75 | 75 | 64 | 69 | 66 | 62 | **7** |
|  |  |  |  |  |  |  | 70 | 59 | 61 | 57 | 49 | **8** |
|  |  |  |  |  |  |  |  | 62 | 62 | 52 | 59 | **9** |
|  |  |  |  |  |  |  |  |  | 48 | 49 | 41 | **10** |
|  |  |  |  |  |  |  |  |  |  | 48 | 41 | **11** |
|  |  |  |  |  |  |  |  |  |  |  | 38 | **12** |
|  |  |  |  |  |  |  |  |  |  |  |  | **13** |

## Discussion

We observed the sensitivity of the individual kits ranging from 18%-74%. However the sensitivity improved with time since confirmed by PCR similar to other findings(3).

When we used these kits in combinations of 2 the overall sensitivity improved up to 90% for the best combination. We provide evidence that use of RDTs in an algorithm may improve their performance in diagnosis for COVID-19.

Further work is required to explore additional RDTS and combinations tested in parallel or series.

## Data Availability

Data is readily Available

